# Association of hypertension, diabetes, stroke, cancer, kidney disease, and high-cholesterol with COVID-19 disease severity and fatality: a systematic review

**DOI:** 10.1101/2020.06.16.20132639

**Authors:** Nazar Zaki, Hany Alashwal, Sahar Ibrahim

## Abstract

**Objective:** To undertake a review and critical appraisal of published/preprint reports that offer methods of determining the effects of hypertension, diabetes, stroke, cancer, kidney issues, and high-cholesterol on COVID-19 disease severity.

**Data sources:** Google Scholar, PubMed, COVID-19 Open Research Dataset: a resource of over 128,000 scholarly articles, including over 59,000 articles with full text related to COVID-19, SARS-CoV-2, and coronaviruses.

**Methods:** A search was conducted by two authors independently on the freely available COVID-19 Open Research Dataset (CORD-19). We developed an automated search engine to screen a total of 59,000 articles in a few seconds. The search engine was built using a retrieval function that ranks a set of documents based on the query terms appearing in each document regardless of their proximity within the document. Filtering of the articles was then undertaken using keywords and questions, e.g. “Effects of diabetes on COVID/normal coronavirus/SARS-CoV-2/nCoV/COVID-19 disease severity, mortality?”. The search terms were repeated for all the comorbidities considered in this paper. Additional articles were retrieved by searching via Google Scholar and PubMed.

**Findings:** A total of 54 articles were considered for a full review. It was observed that diabetes, hypertension, and cholesterol levels possess an apparent relation to COVID-19 severity. Other comorbidities, such as cancer, kidney disease, and stroke, must be further evaluated to determine a strong relationship to the virus. Reports associating cancer, kidney disease, and stroke with COVID-19 should be carefully interpreted, not only because of the size of the samples, but also because patients could be old, have a history of smoking, or have any other clinical condition suggesting that these factors might be associated with the poor COVID-19 outcomes rather than the comorbidity itself. Such reports could lead many oncologists and physicians to change their treatment strategies without solid evidence and recommendations. Further research regarding this relationship and its clinical management is warranted. Additionally, treatment options must be examined further to provide optimal treatment and ensure better outcomes for patients suffering from these comorbidities. It should be noted that, whether definitive measurements exist or not, the care of patients as well as the research involved should be largely prioritized to tackle this deadly pandemic.

## 1. Introduction

The recent outbreak of the novel COVID-19 disease and its rapid worldwide spread poses a global health emergency. The novel virus is thought to belong to the same family as Middle East Respiratory Syndrome (MERS) coronavirus and Severe Acute Respiratory Syndrome (SARS) coronavirus, but it is unique in its own right [10]. Researchers around the globe are investigating several risk factors that may contribute to the severity of COVID-19. The World Health Organization (WHO) indicated that elderly individuals, as well as those with underlying medical conditions, are at higher risk of developing severe COVID-19 disease [11]. As of today, June 14^th^, 2020, more than 7.800 million people are infected by the virus and more than 430,500 have lost their lives. Most of the deaths are believed to be associated with the existence of comorbidities. Generally, patients with compromised immune systems are considered to be particularly vulnerable [12]. Several research works have targeted the epidemiological and clinical characteristics of patients infected with COVID-19; however, the risk factors for severity and mortality have not been sufficiently investigated [13]. Identifying major risk factors and taking corresponding clinical measures could contribute massively to saving lives. Most of the previous studies have identified comorbidities such as hypertension, diabetes mellitus, stroke, cancer, kidney issues, and high cholesterol among the high-risk factors. Thus, this paper represents a systematic review of the literature to shed light on the effects of these pre-existing conditions on COVID-19 disease severity. Additionally, as no method currently exists that can determine the level of risk these comorbidities pose, this paper serves the purpose of achieving the only reasonable option: to draw lessons from earlier experiences and practices.

## 2. Method

A search was conducted on the freely available COVID-19 Open Research Dataset (CORD-19) [14]. CORD-19 is a resource consisting of 128,000 scholarly articles, including over 59,000 articles with full text related to COVID-19, SARS-CoV-2, and coronaviruses. The dataset is frequently updated when new research appears in peer-reviewed publications and archive services became available. To screen every article in the dataset, we developed a search engine in addition to the BM25 search index [15]. BM25 is a retrieval function that ranks a set of documents based on the query terms appearing in each document, regardless of their proximity within the document. All papers are indexed by simply applying a preprocessing function to clean and tokenize the abstracts. Once all documents were indexed, we created document vectors by loading optimized, cached JSON tokens followed by applying a document similarity index using Annoy [16]. Annoy is a C++ library with Python bindings to search for documents in space that are close to a given query. For more details of the implantation, please visit https://github.com/dgunning/cord19.

Filtering of the articles was then undertaken using keywords and questions, e.g. “Effects of diabetes on COVID/normal coronavirus/SARS-CoV-2/nCoV/COVID-19 disease severity, mortality?”. The search terms are repeated for all the comorbidities considered in this paper.

## 3. Results

As illustrated in Fig. 1, a systematic approach was used to screen the full COVID-19 Open Research Dataset [14]. Through the systematic method, 68 articles were found that scored highly on the confidence rating. An additional search was run, employing Google Scholar and PubMed, to ensure every relevant article was included. Each of the 68 papers was read thoroughly, causing 34 articles to be excluded from the list and an additional 19 articles to be manually added to the list. The ultimate list of 54 articles was split into six categories, with 7 articles focusing on cancer, 11 on diabetes mellitus, 7 on kidney issues, 9 on stroke, 12 on hypertension, and 8 on high cholesterol.

**Fig. 1:**
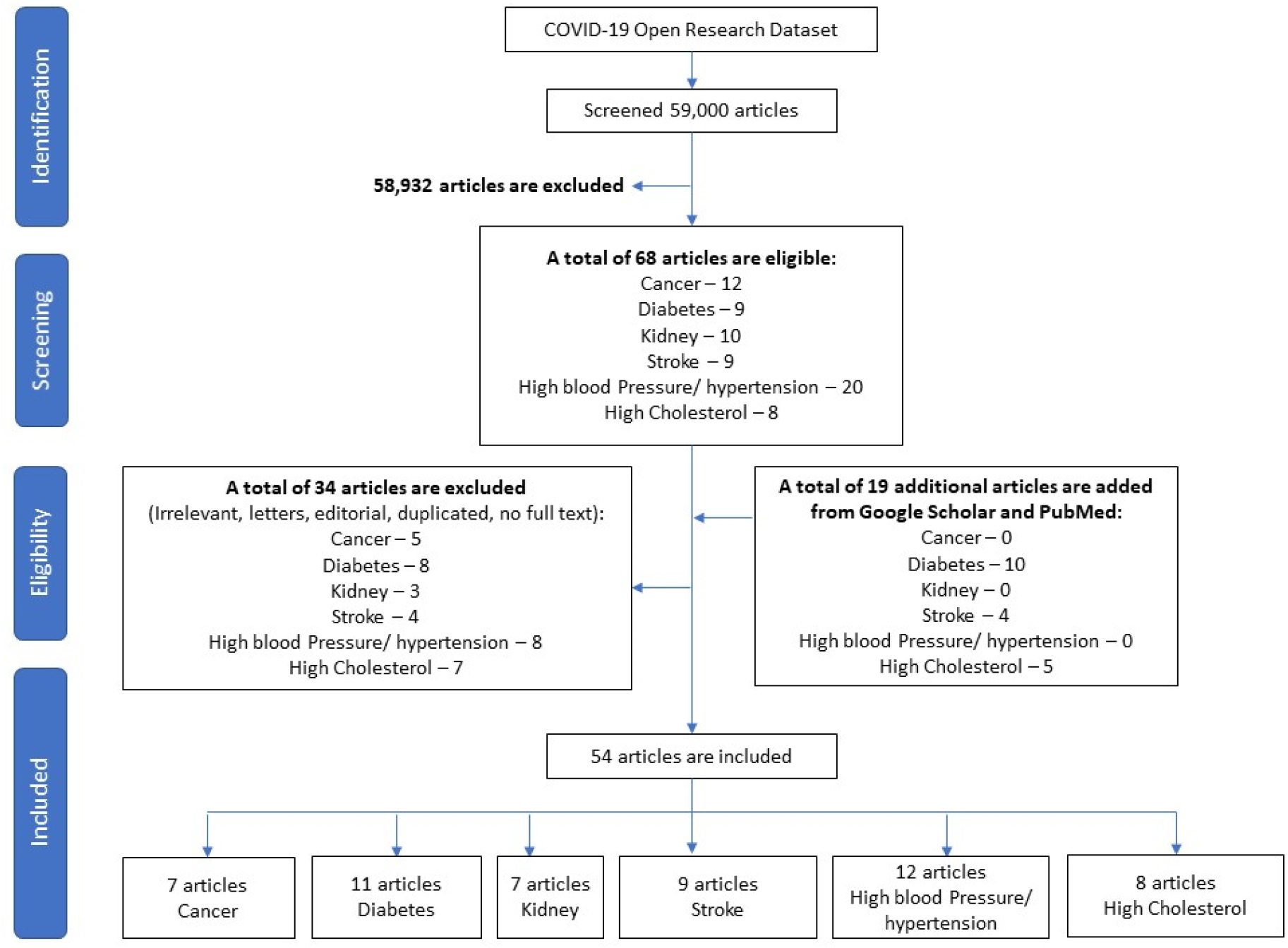
Flowchart of the study inclusions and exclusions of articles

### 3.1. Diabetes

Diabetes is one of the most serious comorbidities linked to the severity of all three well-known human pathogenic coronavirus infections, including SARS-CoV 2. Patients with diabetes have an increased risk of severe complications, including Adult Respiratory Distress Syndrome (ARDS) and multi-organ failure. Depending on the global region, 20–50% of COVID-19 pandemic patients had diabetes, which emphasizes the importance of the link between COVID-19 and diabetes [17]. According to Gupta R. et al. [1], individuals with diabetes are at risk of infections, especially influenza and pneumonia. Patients with diabetes are severely affected when they contract respiratory viruses. Therefore, diabetes was seen as an important risk factor for mortality in patients infected with pandemic influenza A 2009 (H1N1), SARS coronavirus, and Middle East Respiratory Syndrome-related coronavirus (MERSCoV) [18], [19], [20].

Several studies show that among the COVID-19 patients, there was also a noticeable percentage of patients suffering from diabetes mellitus (DM). Yang X. et al. [21] found that among the 32 non-survivors, from a group of 52 COVID-19 patients (median age of 59.7, 67.31% male), 22% were diabetes patients. Additionally, out of 1,099 patients with confirmed COVID-19 (median age of 47, 58.1% male) from autonomous regions in China, 173 had severe disease with comorbidities in which 16.2% were diabetes mellitus [22]. Zhang JJ. et al. [23] analyzed data for 140 patients (median age of 57, 50.7% male) who were admitted to hospitals in Wuhan, China with confirmed COVID-19 cases; 12.1% of these patients had diabetes.

Moreover, Zhang Y. et al. [24] reviewed 258 confirmed COVID-19 patients (63 with diabetes) who were hospitalized at the West Court of Union Hospital in Wuhan, China. The patient median age was 64 and no significant difference was observed between males and females. However, as indicated by the authors, patients with diabetes had significantly higher leukocyte and neutrophil counts as well as higher levels of fasting blood glucose, serum creatinine, urea nitrogen, and creatine kinase isoenzyme MB at admission, compared to those without diabetes. The Cox proportional hazard model showed that diabetes (adjusted hazard ratio [aHR]=3.64; 95% CI: 1.09-12.21) and fasting blood glucose (aHR=1.19; 95% CI: 1.08-1.31) were associated with the fatality of COVID-19, adjusting for potential confounders. The authors concluded that diabetes mellitus is associated with greater disease severity as well as a higher risk of mortality in patients with COVID-19, and recommended that primary and secondary prevention strategies are needed for COVID-19 patients with diabetes.

Bloomgarden ZT. et al. [25] stated that mortality rates from pneumonia among patients with diabetes in Hong Kong (age >= 75) exceed mortality rates in this age group from cardiovascular disease and cancer [26]. Similar evidence of risk amid persons with diabetes has been reported for the two earlier CoV infections: SARS in 2002 (affecting more than 8,000 individuals, mainly in Asia [27], [28]) and MERS in 2012 (affecting more than 2,000 individuals, mainly in Saudi Arabia [29]).

Furthermore, diabetes was present in 42.3% of 26 fatalities due to COVID-19, in a study from Wuhan, China [30]. Huang C. et al. [31] investigated 41 hospital-admitted patients identified as having a laboratory-confirmed 2019-nCoV infection. Most of the infected patients were men (73%), the median age was 49 years and 20% had diabetes. However, these cases are only a small proportion of the patients affected. A report of 72,314 cases (with 44,672 confirmed COVID-19 cases) published by the Chinese Centre for Disease Control and Prevention showed an increased mortality rate in people with diabetes (2.3% overall and 7.3% in patients with diabetes) [32].

Bello-Chavolla OY. [33] predicted mortality due to SARS-CoV-2 and developed a mechanistic score 2 relating obesity and diabetes to COVID-19 outcomes, in data from Mexico. Among 71,103 subjects (predominantly male, the median age of 56.9), the authors observed 15,529 subjects with SARS-CoV-2 and 1,434 deaths (from SARS-CoV-2 or other causes). Risk factors for lethality in COVID-19 included early-onset diabetes, obesity, advanced age, immunosuppression, and CKD. Additionally, it was observed that obesity mediates 45.5% of the effect of diabetes on COVID-19 lethality. The authors used stratified points according to categories of Low risk (≤0), Mild risk (1-3), 141 Moderate risks (4-6), High risk (7-9), and Very High risk (≥10); the score of diabetes was 1. Out of the 137 deaths from SARS-CoV-2, 2.76% had diabetes.

A total of 174 patients (median age of 59, 43.7% male) confirmed with COVID-19 were studied by Guo W. et al. [34]. Demographic data, medical history, symptoms, signs, laboratory findings, chest computed tomographies (CT), as well as the treatment measures, were collected and analyzed. The authors found that 24 COVID-19 patients with no other comorbidities except diabetes were at higher risk of severe pneumonia, the release of tissue injury-related enzymes, excessive uncontrolled inflammation responses, and hypercoagulable state associated with dysregulation of glucose metabolism. The authors recommended that diabetes should be considered as a risk factor for rapid progression and poor prognosis of COVID-19. Extensive care should be administered to patients with diabetes in case of rapid deterioration. The authors described a mortality rate of COVID-19 amongst diabetic patients, without other comorbidities, of about 16%. The above-mentioned studies are summarized in Table 1.

**Table 1:** The summary of the cases reported on the effects of diabetes on COVID-19 patients

| Ref. | No. of Patients | % of DM | City/Country | Median Age | % Male/Female | Symptoms | Duration | Note |
| --- | --- | --- | --- | --- | --- | --- | --- | --- |
| [21] | 52 | 22.0 | Wuhan, China | 59.7 | 67.3/ 32.7 | Fever-98.1% | Dec 2019, and Jan 26, 2020 | The correlation between DM and COVID-19 was not mentioned |
| [22] | 1,099 | 16.2 | Autonomous regions, China | 47.0 | 58.1/41.9 | Fever-43.8%, Cough-67.8%, Diarrhea- 3.8% | Dec 11, 2019, and Jan 29, 2020 | The correlation between DM and COVID-19 was not mentioned |
| [23] | 140 | 12.1 | Wuhan, China | 57.0 | 50.7/49.3 | Fever- 91.7%, Cough-75.0%, Fatigue-75.0%, Diarrhea- 12.9%, Gastrointestinal-39.6% | NA* | The prevalence of diabetes in China is 10.9% in adults, which was slightly lower than the 12.1%; this may be due to the large ratio of elder COVID-19 patients in this study. |
| [24] | 258 | 24.52 | Wuhan, China | 64.0 | No significant difference | Fever-82.2%, Cough-67.1%, Polypnea-48.1%, Fatigue-38% | Jan 29, 2020, to Feb 12, 2020 | (Pre-print) DM is associated with greater disease severity and a higher risk of mortality in patients with COVID-19. |
| [30] | 26 | 42.3 | Wuhan, China | NA* | NA* | Fever (80.5%), cough (56.1%), short of breath (31.7%), chest tightness/pain (24.4%), fatigue (22.0%), dyspnea (12.2%), dizziness/headache (4.9%), general pain (7.3%) and chills (4.9%) | NA* | Diabetes, along with other comorbidities were found to be dangerous factors that resulted in death. |
| [31] | 41 | 20.0 | Wuhan, China | 49.0 | 73.0/27.0 | Pneumonia-100%, Fever-98%, Cough-76%, Lymphopenia-63%, Dyspnea-55%, Fatigue-44%, Sputum production-28%, Headache-8%, Hemoptysis-5%, Diarrhea- 3% | Jan 2, 2020 | Less than half of the infected patients had underlying diseases (13 [32%]), including diabetes (eight [20%]), hypertension (six [15%]), and cardiovascular disease (six [15%]). |
| [32] | 44,672 | 7.3 | Hubei Province (75%) | 87% between 30 to 79 years | NA* | NA* | Feb 11, 2020 | The correlation between DM and COVID-19 was not mentioned |
| [33] | 137 | 2.76 | Mexico | 56.9 | Predominantly male | NA* | April 27th, 2020 | (Pre-print) The authors used stratified points according to categories of Low risk ($\leq 0$), Mild risk (1-3), 141 Moderate risks (4-6), High risk (7-9), and Very High risk ($\geq 10$); the point of diabetes was 1. |
| [34] | 174 | 13.79 | Wuhan, China | 59 | 43.7/56.3 | Fever (78.2%), chill (68.4%), cough (32.2%), fatigue (27%), chest tightness (25.9%), shortness of breath (24.1%) and myalgia (20.7%) | 10 Feb 2020 to 29 Feb, 2020 | Diabetes should be considered as a risk factor for rapid progression and poor prognosis of COVID-19. |

NA\* - Not available

### 3.2. Cancer

According to a few recently published results (mainly Chinese cohort), patients with cancer are more vulnerable to COVID-19 complications. However, how susceptible cancer patients are to infection with COVID-19 has yet to be established.

Liang W. et al. [6] collected and analyzed 2,007 cases from 575 hospitals, in 31 provincial administrative regions in China. All cases were diagnosed with laboratory-confirmed COVID-19. A total of 417 cases were excluded because of insufficient records of previous disease history. The number of COVID-19 cases with a history of cancer is 18 (1%; 95% CI: 0.61–1.65) out of 1,590 COVID-19 cases, which according to 2015 cancer epidemiology statistics seems to be higher than the incidence of cancer in the overall Chinese population (285.83 [0·29%] per 100,000 people. Lung cancer was the most frequent type (five [28%] of the 18 patients). Compared with patients without cancer, patients with cancer were older (mean age 63.1 years), had no significant differences in sex, other baseline symptoms, other comorbidities, or baseline severity of X-ray. Most importantly, patients with cancer were observed to have a higher risk of severe events compared to patients without cancer (seven [39%] of 18 patients vs 124 [8%] of 1,572 patients; Fisher’s exact p=0.0003). The authors concluded that patients with cancer might have a higher risk of COVID-19 than individuals without cancer. Additionally, they showed that patients with cancer had poorer outcomes from COVID-19, providing a timely reminder to physicians that cancer patients require intensive care while infected.

Zhang L., et al. [38] regarded cancer patients as a highly vulnerable group in the current COVID-19 pandemic. The authors included 28 cancer patients with laboratory-confirmed COVID-19, from 3 designated hospitals in Wuhan, China, from Jan. 2020 to 26 Feb. 2020. The median age of the patients was 65, 60.7% were males, and 25% had lung cancer. The authors mentioned that cancer patients show deteriorating conditions and poor outcomes from the COVID-19 infection.

In addition, Yu J. et al. [39] reviewed the medical records, including demographic, clinical, and treatment data, of 1,524 patients with cancer who were admitted to Zhongnan Hospital of Wuhan University, China, from Dec. 30, 2019, to Feb. 17, 2020. Outcomes of COVID-19 among patients with cancer were reported. The authors estimated the infection rate of SARS-CoV-2 in patients with cancer, from a single institution, to be 0.79% (12 of 1524 patients: 95% CI: 0.3-1.2). This was higher than the cumulative incidence of all diagnosed COVID-19 cases that were reported in the city of Wuhan over the same time (0.37%; 41 152 of 11 081 000 cases; data cutoff on February 17, 2020). The median age of infected patients was 66 years. Seven of 12 (58.3%) patients had non–small cell lung carcinoma (NSCLC). Five (41.7%) were being treated with either chemotherapy with or without immunotherapy (n = 3) or radiotherapy (n = 2). Three patients (25.0%) developed SARS; and one patient required intensive-level care. By March 10, 2020, 6 patients (50.0%) were discharged, whereas 3 deaths (25.0%) were recorded. Of the 1,524 patients with cancer who were screened, 228 had NSCLC. The authors stated that patients with NSCLC older than 60 years had a higher incidence of COVID-19 than those aged 60 years or younger (4.3% vs 1.8%). In Table 2, we summarize the details of the three mentioned case studies.

**Table 2:** The summary of the cases reported on the effects of cancer on COVID-19 patients

| Ref. | Cases | Age | Types | M/F | symptoms | Severity | Mortality Rate | Note |
| --- | --- | --- | --- | --- | --- | --- | --- | --- |
| [6] | 18 | Mean = 63.1 | Lung – 5 (27.78%) | No significant difference | No significant difference | No significant difference | NA* | - Postponed adjuvant chemotherapy or elective surgery for stable cancer in endemic areas.<br>- Stronger personal protection provisions for cancer patients.<br>- Intensive surveillance/ treatment for cancer patients with COVID-19, especially in older patients or those with other comorbidities |
| [38] | 28 | Median = 65.0 | Lung - 7(25%) | 17/11 (60.71%/ 39.29%) | Fever (82.1%), dry cough (81%), and dyspnoea (50.0%), along with lymphopaenia (82.1%), high level of high-sensitivity C-reactive protein (82.1%), anaemia (75.0%), and hypoproteinaemia (89.3%). | 53.6% | 28.6% | - Cancer patients show deteriorating conditions and poor outcomes from the COVID-19 infection. |
| [39] | 12 | Median = 66 | Lung – 7 (58.33%) | 10/2 83.33%/ 16.67%) | Fever (100%), Dyspnea (25%), Cough (25%), | 25% | 25% | - Patients with cancer from the epicenter of a viral epidemic harbored a higher risk of SARSCoV-2 infection (OR, 2.31; 95% CI: 1.89-3.02) compared with the community.<br>- Older patients (>60 years) and patients with NSCLC may be at risk of COVID-19.<br>- Larger sample sizes in patients with cancer will resolve these potential associations. |

NA\* - Not available

### 3.3. High-Blood Pressure and Hypertension

High blood pressure (hypertension) is one of the most common conditions in severe COVID-19 patients [8]. In this section, we review several studies that looked at the relationship between hypertension and the severity of COVID-19. The relationship between high blood pressure and increased risk of pneumonia was a subject of a study that analyzed UK Biobank data of 107,310 hypertension patients, with 3% of patients developing pneumonia afterward [9]. The data analysis shows that the risk of respiratory disease is significantly higher in patients with hypertension. Patients with hypertension were also found to be at higher risk of acute respiratory disease and chronic lower respiratory disease, independent of age, sex, smoking status, and BMI. Generalizing these results to COVID-19 is plausible and led to many studies that focus on hypertension as a strong indication of COVID-19 severity.

Several recent studies have investigated the variability in COVID-19 disease severity in patients with hypertension and its medicinal therapy. In [40], the authors reported that among 3,017 hospitalized COVID-19 patients, 53% were diagnosed with hypertension. Furthermore, the mortality rates among patients on angiotensin-converting enzyme inhibitors (ACEI) for hypertension treatment (27%, p=0.001) as well as for patients on Angiotensin II receptor blockers (ARBs) (33%, p=0.12) were lower compared to other anti-hypertensive agents (39%). Therefore, these results support the continuation of ACEI and ARB therapy for COVID-19 hypertensive patients.

These results are supported by another study [41], where 126 COVID-19 patients with hypertension were retrospectively grouped into ARBs/ACEIs (n=43) and non-ARBs/ACEIs (n=83) according to their antihypertensive medications. A group of COVID-19 patients without hypertension were then randomly selected to match the previous group based on age and sex. The results of this study show that the ARBs/ACEIs group had a lower proportion of critical patients (9.3% vs 22.9%; p=0.061), and a lower death rate (4.7% vs 13.3%; p=0.216). However, these differences are not statistically significant.

Similarly, Liu et al. [42], investigated the disease severity in patients with hypertension who used ACEI, ARB, CCB, and beta-blockers (BB) compared to patients who did not take any hypertension medication. Considering only 46 elderly patients (age > 65), the results show that patients who took ARB have a significantly lower risk of developing a severe disease compared to patients who took no drugs. However, although patients who took ACEI, CCB, and BB also had a lower risk of developing severe disease, there was no statistical significance compared to patients who took no drugs. This could be due to the limited number of patients on these drugs compared to ARB.

In another study [43], the authors argued, based on their retrospective investigation of different hypertension drugs, that calcium channel blockers (CCBs) show significant efficacy on COVID-19 patients. They also conducted in vitro experiments to examine the efficacy of CCBs and other hypertension drugs on blocking SARS-CoV-2 replication in cells. The results show that only CCBs block virus replication. However, the blocking mechanism is not apparent. Therefore, further investigations of CCBs efficacy on post-entry virus replication in vitro and clinically are needed.

Although the studies cited above show hypertension medications are beneficial for COVID-19 patients, a recent retrospective, observational study [44] found that patients with hypertension, who took ACEI/ARB drugs, have a higher risk of developing a severe case of COVID-19 (P = 0.064). However, this is not a statistically significant difference and may be due to the limited number of patients with hypertension (75 out of 274 COVID-19 patients). Also, the group of patients with hypertension were older and had more comorbidities, such as chronic renal insufficiency, cardiovascular disease, diabetes mellitus, and cerebrovascular disease, than patients without hypertension.

Interestingly, another study [45] hypothesizes that hypertension treatment with ACEI and ARBs, which results in an upregulation of ACE2, may increase the risk of developing severe COVID-19. This is based on a structural study suggesting that SARS-CoV-2 uses ACE2 as its receptor [46]. It was also noted that CCB treatment of hypertension does not lead to an upregulation of ACE2 which in turn makes it a safe treatment of hypertension in COVID-19 patients. However, this theory is not supported by the previously cited retrospective studies on COVID-19 patients who took ACEI and ARBs [40]. Besides, Sanchis-Gomar et al. [47], noted that several health organizations have recommended the continued use of ACEIs and ARBs for treating hypertension in COVID-19 patients. They also found, based on their review study, that there are no significant differences between ACEIs and ARBs for managing hypertension, with ARBs being a more favorable option for severe cases of COVID-19.

We also reviewed several articles that employed systematic reviews and meta-analysis of published studies on the effects of hypertension on COVID-19 disease severity. In [48], systematic reviews show that hypertension is one of the major comorbidities of fatality COVID-19 cases. Similarly, Zuin et al. [49] carried out a systematic review and meta-analysis that show COVID-19 patients with hypertension have a significantly higher mortality risk. However, it is noted that further studies of other comorbidities either linked or not linked to hypertension are needed. Also, Roncon et al. [50] looked at the rate of ICU admission of COVID-19 patients with hypertension. They found that among the 1,382 patients (mean age = 51.5 years, 57.74% males), patients with hypertension had a significantly increased risk of ICU admission (OR=2.54; 95% CI: 1.83 - 3.54; P<0.0001). The above-mentioned cases are summarized in Table 3.

**Table 3:** An overview of retrospective observational studies investigating hypertension effect on COVID-19 severity.

| Ref. | Location/Approach | Patients Groups | Hypertension (HT)/Treatment Effect | Note |
| --- | --- | --- | --- | --- |
| [40] | New Jersey, USA | 3,017 totals, 1,584 with HT | Mortality significantly increased at 35% vs 13% in patients without HT |  |
| [41] | Wuhan, China | 126 with HT<br>43 on ARBs/ACEIs treatment<br>83 non-ARBs/ACEIs | ARBs/ACEIs have a lower proportion of critical patients (9.3% vs 22.9%; p=0.061), and a lower death rate (4.7% vs 13.3%; p=0.216) | The difference is not statistically significant. |
| [42] | Shenzhen, China | 46 with HT (age > 65) | ARBs significantly reduced the risk of a severe case (p=0.025) | Other treatment lowers the risk but not significantly |
| [43] | Wuhan, China | 90 with HT | CCBs reduced mortality rate (6.8%, n=44) compared to ARBs/ACEIs (41.2%, n=17) | The difference is not statistically significant. |
| [44] | Wuhan, China | 274 totals, 75 with HT | ACEIs/ARBs have a higher risk of severe COVID-19 (P = 0.064). | HT patients are older and had other underlying comorbidities |

### 3.4. High-Cholesterol

Cholesterol level alteration has been found to occur during a viral infection such as (HIV) and hepatitis C virus (HCV) [51]. The mechanism of alteration involves the virus binding to the Scavenger receptor B type 1 (SR-B1) that facilitates the selective uptake of high-density lipoprotein (HDL) cholesterol and other lipid components of receptor-bound HDL particles, including free cholesterol (FC) and triglycerides (TG) [52]. Also, it has been found that membrane cholesterol is an important component to facilitate viral entry into host cells [53]. The role of cholesterol in viral infection was investigated even before the current pandemic of SARS-CoV-2. Ren et al. [54], found that cholesterol depletion from the cellular membrane significantly impaired the efficiency of viral infection. However, this study was carried out in vitro on cells from swine testicles (ST) and baby hamster kidney cells (BHK21).

Wei et al., [52] studied the cholesterol metabolism of COVID-19 patients compared to the normal reference population. In general, they found that FC, HDL, and LDL cholesterol level is significantly low for COVID-19 patients compare to the reference population. TG levels were also lower, but not at a significant level. Moreover, cholesterol metabolism of mild, moderate, severe, and critical COVID-19 patients was also presented and discussed. The levels of HDL-C were significantly lower in patients with the severe and critical disease than in patients with moderate or mild disease.

In another study [55], HDL cholesterol levels along with serum albumin levels were used as predictors for the severity of COVID-19 disease. The study investigated the differences in clinical features between mild and severe COVID-19 patients. Other features studied included: hypoproteinemia, hypoalbuminemia, and decreased ApoA1. Patients were grouped into two types: mild and severe, and those in critical condition were not included. It was also noted that patients with severe disease were significantly older (median age: 58) compared to patients with mild disease (median age: 37). Their results show that patients with severe disease have significantly lower HDL cholesterol. They also observed increased levels of albumin and HDL cholesterol in patients who were at the late stage of recovering from COVID-19.

Another recent study [56], looked at the serum cholesterol level in patients with COVID-19 in Wenzhou, China. The laboratory test results show that patients had significantly low FC, HDL-cholesterol, and LDL-cholesterol levels compared to healthy control. Moreover, they investigated the cholesterol levels in two categories of patients: primary infection cases being those who had been in Wuhan, and secondary infection cases being those who had never been in Wuhan but were infected by person-to-person transmission. They reported that HDL-cholesterol in the primary infection group was significantly lower than in the secondary infection group. Similar to the results in [55], the cholesterol levels of FC, HDL, and LDL were improving as patients recovered.

The inverse correlation between HDL cholesterol level and severity of COVID-19 is also highlighted in [57]. The author argued that taking statin drug therapy for cholesterol may increase COVID-19 infections. Statins are drugs used to lower cholesterol and protect against a heart attack and stroke. Taking statins results in very low or no production of endogenous cholesterol, which leads to the upregulation of LDL in the cell membrane. This, as stated earlier, is found to be an important component to facilitate viral entry into host cells. Overview of articles investigating cholesterol levels in COVID-19 patients is summarized in Table 4.

**Table 4:** An overview of articles investigating cholesterol levels in COVID-19 patients.

| Ref. | Location/Approach | Patients Groups | Cholesterol Level | Note |
| --- | --- | --- | --- | --- |
| [52] | Wuhan, China | 861 patients<br>1,108 normal population | TC, HDL, and LDL significantly lower in patients compared to normal ($p < 0.0001$) | TC, HDL, and LDL, were also significantly lower in critical and severe patients compared to mild patients. |
| [55] | Wuhan, China | 97 patients.<br>Mild (n = 72)<br>Severe group (n = 25) | HDL is significantly lower in severe compared to mild ($p < 0.001$) | TC and LDL were lower in severe patients but not insignificant values |
| [56] | Wenzhou Medical University, China | 71 patients<br>80 controls | TC, HDL, and LDL significantly lower in the patient compared to controls ($p < 0.001$) | Lower cholesterol level in Primary infections (Wuhan patients) compared to secondary infection (person-to-person) ($p < 0.05$) |
| [53] | In vitro cells | NA* | A higher level of cholesterol facilitated the entry of pathogenic viruses. | An important finding is that SARS-CoV had a higher binding affinity to Cholesterol-rich membranes. |
| [54] | In vitro cells | NA* | Virus infection efficiency was reduced by cholesterol depletion and was restored when the concentration of cholesterol was increased | The cholesterol-rich membrane is important for virus entry |
| [57] | Review | NA* | Statin drugs that indirectly increase membrane cholesterol may increase COVID-19 infection | The author presented an argument that commonly prescribed statin drugs in European countries may have contributed to the increased COVID-19 infection |

NA\* - Not available

### 3.5. Kidney Issues

Apart from fever and respiratory complications, acute kidney injury has been observed in some patients with COVID-19. To investigate the possible cause of kidney damage in COVID-19 patients, Lin W. at al. [58] used both published kidney and bladder cell atlas data as well as an independent, unpublished kidney single-cell RNA-Seq data generated in-house, to evaluate ACE2 gene expressions in all cell types of healthy kidneys and bladders. The results showed the enriched expression of all subtypes of proximal tubule cells of the kidney and low but detectable levels of expression in bladder epithelial cells. These results indicated the urinary system is a potential route for COVID-19 infection, along with the respiratory system and digestion system. The findings suggested the kidney abnormalities of SARS and COVID-19 patients may be due to proximal tubule cell damage and subsequent systematic inflammatory response induced kidney injury. Beyond that, laboratory tests of viruses and related indicators in urine may be needed in some special patients of COVID-19.

As for the novel coronavirus, a recent study reported that the human kidney is a specific target for SARS-CoV-2 infection [59]. The authors examined the viral nucleocapsid protein in situ in the kidney post-mortem and found that SARS-CoV-2 antigens accumulated in kidney tubules, suggesting that SARS-CoV-2 infects the human kidney directly, inducing AKI and contributing to viral spreading in the body [60].

Alberici F. et al. [61] describe 20 kidney transplant recipients (median age 59 years, 80% male) with SARS-CoV2 induced pneumonia. All patients presented with fever but only one complained of difficulty in breathing. Overall, 5 kidney transplant recipients died after a median period of 15 days from symptom onset. These preliminary findings describe a rapid clinical deterioration, associated with chest radiographic deterioration and escalating oxygen requirement in renal transplant recipients with SARS-Cov2 pneumonia. Thus, in this limited cohort of long-term kidney transplant patients, SARS-CoV-2 induced pneumonia is characterized by a high risk of progression and significant mortality.

Wang J. et al. [62] conducted a study on one male patient (age 49) with a history of close contact with a confirmed COVID-19 case, type 2 diabetes for 20 years, and hypertension for 10 years. The patient underwent comprehensive treatment for COVID-19. Organ transplant patients with COVID-19 infection might have poorer prognosis because of their systemic immunosuppressive state. However, this severe case was cured even without discontinuing or reducing his immunosuppressant therapy. This immunosuppressed case might help physicians to establish optimal treatment strategies for similar severe cases.

Whether COVID-19 causes significant acute kidney injury (AKI) remains controversial. Zhou H. et al. [63] retrospectively analyzed the clinical characteristics, urine, and blood routine tests as well as other laboratory parameters of 178 hospitalized COVID-19 patients in Wuhan, China. No patient presented increased serum creatinine (Scr), and 5 (2.8%) patients showed increased blood urea nitrogen (BUN), indicating a few cases with “kidney dysfunction”. However, among the 83 (46.6%) patients with no history of kidney disease who received routine urine test upon hospitalization, 45 (54.2%) patients displayed abnormality in urinalyses, such as proteinuria, hematuria, and leukocyturia, and none of the patients were recorded to have acute kidney injury (AKI) throughout the study. Meanwhile, the patients with abnormal urinalysis typically displayed worse disease progression that was reflected through laboratory parameter presentations, including markers of liver injury, inflammation, and coagulation. The authors concluded that urinalysis is better in unveiling potential kidney impairment of COVID-19 patients than blood chemistry tests. Additionally, urinalysis could be used to reflect and predict disease severity.

Cheng Y. et al. [64] determined the prevalence of acute kidney injury (AKI) in patients with COVID-19 and evaluated the association between markers of abnormal kidney function and death in patients with COVID-19. The authors studied a cohort of 701 patients (median age of 63, 52.35% male) with COVID-19, in Wuhan during 2020, of whom 113 (16.1%) died in hospitals. During the study period, AKI occurred in 5.1% of patients. Kaplan-Meier analysis demonstrated that patients with kidney disease had a significantly higher risk of in-hospital death. Thus, the authors believed that the prevalence of kidney disease on admission and the development of AKI during hospitalization in patients with COVID-19 is high and is associated with in-hospital mortality. Hence, clinicians should increase their awareness of kidney disease in patients with severe COVID-19.

### 3.6. History of Stroke

Acute stroke remains a medical emergency even during the COVID-19 pandemic. Avula A. et al. [65] investigated 4 patients with COVID-19 that also presented acute stroke. All 4 patients (Table 5) had radiographic evidence of stroke and PCR-confirmed COVID-19 infection. The authors elucidated the clinical characteristics, imaging findings, and the clinical course. They warned that COVID-19 patients can present with cerebrovascular accidents and should use appropriate personal protective equipment in every suspected patient. Further studies are urgently needed to improve the current understandings of neurological pathology in the setting of COVID-19 infection.

**Table 5:** Details of the 4 patients that had radiographic evidence of stroke and PCR-confirmed COVID-19 infection

| Patient | Age | Gender | Past Medical History | Symptoms | Treatment | Note |
| --- | --- | --- | --- | --- | --- | --- |
| 1 | 73 | M | Hypertension, dyslipidemia, and carotid stenosis | Fever, respiratory distress, and altered mental status | Aspirin and other supportive measures | The family chose for terminal extubation with comfort measures. |
| 2 | 83 | F | Frequent urinary tract infections, hypertension, hyperlipidemia, diabetes mellitus type 2, and neuropathy | Fever, facial droop, slurred speech, and reduced oral intake | NA* | The family chose for terminal extubation with comfort measures. |
| 3 | 83 | F | Hypertension | Mental issues and left-sided weakness | NA* | The family chose for terminal extubation with comfort measures. |
| 4 | 88 | F | Hypertension, chronic kidney disease, and hyperlipidemia | The transient 15-minute episode of right arm weakness and numbness along with word-finding difficulty. Mild shortness of breath with dry cough | Aspirin, statins | NA* |

NA\* - Not available

Klok et al. [66] evaluated the incidence of the composite outcome of ischemic stroke in 184 patients admitted to the ICU of 3 Dutch hospitals. The patients were proven to have COVID-19 induced pneumonia, of whom 23 died (13%). All patients received at least the standard doses of thromboprophylaxis. The cumulative incidence of the composite outcome was 31% (95%CI 20-41). The 31% incidence of thrombotic complications in ICU patients with COVID-19 infections is remarkably high. Therefore, the authors recommended to strictly apply pharmacological thrombosis prophylaxis in all COVID-19 patients admitted to the ICU.

Lodigiani C. et al. [67] studied 388 symptomatic patients (median age 66 years, 68% men, 16% requiring ICU admittance with laboratory-proven COVID-19) admitted to a university hospital in Milan, Italy (from Feb 13, 2020, to April 10, 2020). The primary outcome was a thromboembolic complication, including venous thromboembolism (VTE), ischemic stroke, and acute coronary syndrome (ACS)/myocardial infarction (MI). The secondary outcome was overt disseminated intravascular coagulation (DIC). Thromboembolic events occurred in 28 patients (7.7% of closed cases; 95%CI 5.4%–11.0%), corresponding to a cumulative rate of 21% (27.6% ICU, 6.6% general ward). The high number of arterial and, in particular, venous thromboembolic events diagnosed within 24 hours of admission as well as the high rate of positive VTE imaging tests (among the few COVID-19 patients tested), suggest that there is an urgent need to improve specific VTE diagnostic strategies as well as investigate the efficacy and safety of thromboprophylaxis in ambulatory COVID-19 patients.

Mao L. et al. [68] studied the neurologic manifestations of patients with COVID-19 in Wuhan, China. The authors collected data from Jan 16, 2020, to Feb 19, 2020, at 3 designated special care centers for COVID-19 of the Union Hospital of Huazhong University of Science and Technology in Wuhan, China. The study included 214 (mean age 72.7 years, 40.7% men, 58.9% had a non-severe infection and 88 patients (41.1%) had a severe infection) consecutive hospitalized patient with a laboratory-confirmed diagnosis of COVID-19. However, it has not been reported that patients with COVID-19 had any neurologic manifestations.

In a note to Eur. Neurol journal editor, Morelli N. et al. [69] mentioned that between February 21, 2020 (first COVID-19 patient recorded in Italy), and March 25, 2020, there were only 6 admissions from the Casualty Department for ischemic stroke. The authors suggested that the reason could be the controversial role IL-6 plays in stroke. Although high IL-6 levels have been reported to harm brain infarct volume and long-term outcome [70], in ischemic stroke there has also been experimental evidence that IL-6 has a protective effect and helps in the improvement of poststroke angiogenesis [71]. This could also be due to the presence of thrombocytopenia in COVID-19 patients. Indeed, what may be true for influenza pneumonia (i.e., increased stroke risk) may not be true for SARSCoV-2. The main limitation of the remark, however, is certainly the short observation period of just 1 month.

Oxley TJ. et al. [72] reported 5 (4 male, 1 female) large-vessel stroke patients with confirmed SARS-CoV-2 (mean age is 40.4, all patients are younger than 50 years) who presented in the health system of New York City. Severe acute respiratory syndrome coronavirus 2 (SARS-CoV-2) infection was diagnosed in all five patients. Although the patients were having COVID-19 related symptoms such as fever, cough, and lethargy, all of them had a reduced NIH Stroke Scale/Score (NIHSS).

## 4. Discussion and Conclusion

The studies reviewed in this paper have assessed the effects of diabetes, cancer, hypertension, high cholesterol, kidney issues, and stroke, on COVID-19 severity. Through a thorough examination of the evidence provided in these reports, inferences on the relationship between these preexisting conditions and COVID-19 can be made.it is apparent that individuals with diabetes are at an increased risk for COVID-19 infection and medical complications including death. Suggestions are made on the possible pathophysiological mechanisms of the relationship between diabetes and COVID-19, and its management. No definite conclusions can be made based on current limited evidence and data [1]. Further research regarding this relationship and its clinical management is warranted [2].

Furthermore, the article by Guo et al. [3] has left several relevant questions to be addressed: Does the association between COVID-19 and diabetes differ by different types of diabetes? Do antidiabetic drugs have an impact on disease progression? Is the impact of aging the same in diabetics as is in those without diabetes? While the results of this study should be read in the light of some limitations, such as the small sample size and the large age difference between study groups when patients with other comorbidities were excluded, it still provides relevant insights that could inform about how COVID-19 interacts with preexisting conditions [4].

A few recently published reports provided results suggesting that cancer patients are susceptible to COVID-19 complications. In these studies, lung cancer was most frequently observed, and the COVID-19 cases analyzed were all from hospitals in China. However, according to Moujaess E. et al. [5], such reports could lead many oncologists to change their daily practice in cancer care, without solid evidence and recommendations. The data presented should be carefully interpreted, not only because of the small sample but also because cancer patients in this cohort (mentioned in [6] for instance) had a significantly older median age than their control (63·1 vs 48·7 years) as well as a more significant history of smoking, suggesting that these two factors might be associated with the poor COVID-19 outcomes than the cancer history itself. Moreover, these results should be interpreted with caution in the absence of adjustment to the prevalence of the different comorbidities in the Chinese population [5]. Also, Xia Y., et al. [7] commented on the mentioned studies and highlighted reasons to be cautious, suggesting that current evidence remains insufficient to explain a conclusive association between cancer and COVID-19. Though one effect that has risen concern, is due to the number of COVID-19 patients surpassing hospital and healthcare capacity, cancer patients may be overlooked resulting in negative consequences in their treatments.

Hypertension, like diabetes, has been proven to be linked to a significantly higher risk of respiratory infection, making it a solid indicator of COVID-19 severity. Additionally, systematic reviews indicate hypertension to be common comorbidity in COVID-19 death cases as well as be linked to significantly higher mortality risk and increased incidence of ICU admission. Although, it should be noted that further studies of other comorbidities connected to hypertension are necessary. Nonetheless, it is evident that many patients with the virus also suffer from hypertension and as a result, the comorbidity as well as its medicinal therapies on COVID-19 have been observed. It is also found that hypertension treatment should be continued to lower the severity risk. Some researchers recommended CCBs treatment and others found there are no significant differences in using ACEIs, ARBs, or CCBs. However, there seems to be a general agreement with most health organizations, who recommended hypertension treatment to be continued in COVID-19 patients [47].

Cholesterol has previously been investigated concerning many viral infections. Typically, there is a selective uptake of cholesterol as viruses bind to the cellular membrane, which ultimately enables severe infection. Therefore, analyses have been conducted on the connection between COVID-19 and cholesterol. Most publications indicated that FC, HDL, and LDL cholesterol levels were significantly low in patients with a viral infection, compared to the control. These studies explained the ability of SARS-CoV-2 virus to use serum cholesterol for its entry into host cells. Therefore, low serum cholesterol level can be used as an early predictor of COVID-19 severity in patients.

Also, observing improvement in serum cholesterol levels may indicate the improvement of conditions in patients. It is also worth noting that statin drugs for cholesterol may contribute to the severity of COVID-19 disease by inhibiting endogenous cholesterol, leading to the upregulation of LDL. This leads to higher membrane cholesterol levels that enhance the ability of SARS-CoV-2 to enter host cells.

In addition to the multiple complications observed, acute kidney injury has also been detected in COVID-19 patients. Results indicated the urinary system, respiratory system, and digestion system as potential routes for the COVID-19 infection; one study indicating the human kidney to be a specific target of the virus. Findings propose kidney abnormalities in COVID-19 patients may be due to proximal tubule cell damage and subsequent systematic inflammatory response induced kidney injury. Additionally, in one study, SARS-CoV-2 antigens were seen to accumulate in kidney tubules, implying the direct impact on the kidney, leading to AKI induction and viral spread within the body. There have also been studies describing a rapid clinical deterioration, escalating oxygen requirement, high risk of progression, and significant mortality in kidney transplant patients. Similarly, the incidence of kidney disease on admission as well as the development of AKI is correlated with fatality in COVID-19 patients. However, if COVID-19 causes acute kidney injury remains uncertain. Many authors concluded that clinicians must still try to increase their awareness of kidney issues in patients with severe infection, and a urinalysis may be used to predict the extent of disease severity.

One study identified 36.4% of COVID patients to have neurological symptoms (mainly in patients with severe disease) and several mechanisms of COVID-19 that increase stroke risk have been recognized but neither has been proven as significant. Further studies are required to investigate the neurological pathology during COVID-19 infection. Additionally, there may be a need to improve specific VTE diagnostic strategies, the safety of thromboprophylaxis, and further identify the role IL-6 plays in ischemic stroke. However, the risks associated with ischemic stroke in COVID-19 patients are still uncertain. As with cancer patients, medical resources must be allocated to target the strain COVID-19 has placed on ischemic stroke care. Meanwhile, the relationship between COVID-19 and stroke must be examined further.

## Data Availability

This is a review article.

## Acknowledgment

The authors would like to thank Mr. Dwight Gunning for his kind assistance with the search engine.

## References

[1] Zaki N., Mohamed EA., “The estimations of the COVID-19 incubation period: a systematic review of the literature,” medRxiv, 2020.

[2] WHO, “ Coronavirus disease (COVID-19) Pandemic,” World Health Organization, 2020.

[3] Burki TK., “Cancer care in the time of COVID-19,” The Lancet, vol. 21, 2020.

[4] Zhou F., Yu T., Du R., Fan G., Liu Y., et al., “Clinical course and risk factors for mortality of adult inpatients with COVID-19 in Wuhan, China: a retrospective cohort study,” The Lancet, p. 1054–1062, 2020.

[5] Wang LL. and Lo K., Chandrasekhar Y., Reas R., Yang J. etal., “CORD-19: The Covid-19 Open Research Dataset,” ArXiv, vol. abs/2004.10706, 2020.

[6] Manning CD., Raghavan P., Schütze H., An Introduction to Information Retrieval, Cambridge, UK: Cambridge University Press, 2009.

[7] Bernhardsson E., “Annoy,” [Online]. Available: https://github.com/spotify/annoy.

[8] Bornstein SR., Rubino F., Khunti K., Mingrone G., Hopkins D., et al., “Practical recommendations for the management of diabetes in patients with COVID-19,” The Lancet, 2020.

[9] Gupta R., Ghosh A., Kumar AS., Misra A., “Clinical considerations for patients with diabetes in times of COVID-19 epidemic,” Diabetes & Metabolic Syndrome: Clinical Research & Reviews, vol. 14, no. 3, pp. 211–212, 2020.

[10] Yang JK., Feng Y., Yuan MY., Yuan SY., Fu HJ. et al., “ChanPlasma glucose levels and diabetes are independent predictors for mortality and morbidity in patients with SARS,” Diabet Med, vol. 23, no. 6, pp. 623–628, 2006.

[11] Schoen K., Horvat N., Guerreiro NFC., Giassiks I., “Spectrum of clinical and radiographic findings in patients with diagnosis of H1N1 and correlation with clinical severity,” BMC Infect Dis., vol. 19, no. 1, p. 964, 2019.

[12] Song Z., Xu Y., Bao L., Zhang L., Yu P. et al., “QinFrom SARS to MERS, thrusting coronaviruses into the spotlight,” Viruses, vol. 1, p. 11, 2019.

[13] Yang X., Yu Y., Xu J., et al., “Clinical course and outcomes of critically ill patients with SARS-CoV-2 pneumonia in Wuhan, China: a single-centered, retrospective, observational study,” Lancet Respir Med, vol. 2600, no. 20, 2020.

[14] Guan W., Ni Z., Hu Y., et al., “Clinical characteristics of coronavirus disease 2019 in China,” N Engl J Med, 2020.

[15] Zhang JJ., Dong X., Cao YY., et al., “Clinical characteristics of 140 patients infected by SARS-CoV-2 in Wuhan, China,” Allergy, 2020.

[16] Zhang Y., Cui Y., Shen M., Zhang J., Liu B., “Comorbid Diabetes Mellitus was Associated with Poorer Prognosis in Patients with COVID-19: A Retrospective Cohort Study,” medRxiv, 2020.

[17] Bloomgarden ZT., “Diabetes and COVID-19,” Journal of Diabetes, vol. 12, p. 347–349, 2020.

[18] Wu H., Lau ESH., Ma RCW., et al., “Secular trends in all-cause and cause-specific mortality rates in people with diabetes in Hong Kong 2001-2016: a retrospective cohort study,” Diabetologia, 2020.

[19] Huang YT., Lee YC., Hsiao CJ., “Hospitalization for ambulatorycare-sensitive conditions in Taiwan following the SARS outbreak: a population-based interrupted time series study,” J Formos Med Assoc., vol. 108, pp. 386–394, 2009.

[20] Chan-Yeung M., Xu RH., “SARS: epidemiology,” Respirology, vol. 8, pp. S9–S14, 2003.

[21] Morra ME., Van Thanh L., Kamel MG., et al., “Clinical outcomes of current medical approaches for Middle East respiratory syndrome: a systematic review and meta-analysis,” Rev Med Virol., vol. 28, p. e1977, 2018.

[22] Deng SQ., Peng HJ., “Characteristics of and public health responses to the coronavirus disease 2019 outbreak in China,” J Clin Med, vol. 2, no. E575, p. 9, 2020.

[23] Huang C., Wang Y., Li X., Ren L., Zhao J., “Clinical features of patients infected with 2019 novel coronavirus in Wuhan, China,” The Lancet, vol. 395, 2020.

[24] Wu Z., McGoogan JM., “Characteristics of and important lessons from the coronavirus disease 2019 (COVID-19) outbreak in China: summary of a report of 72 314 cases from the Chinese center for disease control and prevention,” J Am Med Assoc., 2020.

[25] Bello-Chavolla OY., Bahena-López JP., AntonioVilla NE., Vargas-Vázquez A., González-Díaz A., et al., “Predicting mortality due to SARS-CoV-2: A mechanistic score 2 relating obesity and diabetes to COVID-19 outcomes in Mexico,” MedRxiv, 2020.

[26] Guo W., Li M., Dong Y., Zhou H., Zhang Z., et al., “Diabetes is a risk factor for the progression and prognosis of COVID-19,” Diabetes Metabolism Research and Reviews, vol. 31, p. e3319, 2020.

[27] Liang W., Guan W., Chen R., Wang W., Li J., Xu K., et al., “Cancer patients in SARS-CoV-2 infection: a nationwide analysis in China,” Lancet Oncol., vol. 21, no. 3, pp. 335–337, 2020.

[28] Zhang L., Zhu F., Xie L., Wang C., Wang J., et al., “Clinical characteristics of COVID-19-infected cancer patients: a retrospective case study in three hospitals within Wuhan, China,” Annals of Oncology, 2020.

[29] Yu J., Ouyang W., Chua MLK., Xie C., “SARS-CoV-2 Transmission in Patients With Cancer at a Tertiary Care Hospital in Wuhan, China,” JAMA Oncology, 2020.

[30] Guan W., Liang W., Zhao Y., et al., “Comorbidity and its impact on 1590 patients with COVID-19 in China: a nationwide analysis,” European Respiratory Journal, vol. 55, pp. DOI: https://doi.org/10.1183/13993003.00547-2020, 2020.

[31] Zekavat SM., Honigberg M., Pirruccello J., Kohli P., Karlson EW., et al., “Influence of blood pressure on pneumonia risk: Epidemiological association and Mendelian randomisation in the UK Biobank,” medRxiv, p. doi: https://doi.org/10.1101/2020.04.19.20071936, 2020.

[32] Andrew Ip, Parikh K., Parrillo JE., Mathura S., Hansen E., et al., “Hypertension and Renin–Angiotensin–Aldosterone System Inhibitors in Patients with Covid-19,” medRxiv, p. doi: https://doi.org/10.1101/2020.04.24.20077388, 2020.

[33] Yang G., Tan Z., Zhou L., et al., “Angiotensin II Receptor Blockers and Angiotensin-Converting Enzyme Inhibitors Usage is Associated with Improved Inflammatory Status and Clinical Outcomes in COVID-19 Patients With Hypertension,” medRxiv, p. doi: https://doi.org/10.1101/2020.03.31.20038935, 2020.

[34] Liu Y., Huang F., Xu J., et al., “Anti-hypertensive Angiotensin II receptor blockers associated to mitigation of disease severity in elderly COVID-19 patients,” medRxiv, p. doi: https://doi.org/10.1101/2020.03.20.20039586, 2020.

[35] Zhang L., Sun Y., Zeng H., et al., “Calcium channel blocker amlodipine besylate is associated with reduced case fatality rate of COVID-19 patients with hypertension,” medRxiv, p. doi: https://doi.org/10.1101/2020.04.08.20047134, 2020.

[36] Zeng Z., Sha T., Zhang Y., et al., “Hypertension in patients hospitalized with COVID-19 in Wuhan, China: A single-center retrospective observational study,” medRxiv, p. doi: https://doi.org/10.1101/2020.04.06.20054825, 2020.

[37] Fang L., Karakiulakis G., Roth M., “Are patients with hypertension and diabetes mellitus at increased risk for COVID-19 infection?,” The Lancet, vol. 8, pp. doi: https://doi.org/10.1016/S2213-2600(20)30116-8, 2020.

[38] Wan Y., Shang J., Graham R., Baric RS., Li F., “Receptor Recognition by the Novel Coronavirus from Wuhan: an Analysis Based on Decade-Long Structural Studies of SARS Coronavirus,” Journal of Virology, vol. 94, no. 7, pp. doi: https://doi.org/10.1128/JVI.00127-20, 2020.

[39] Sanchis-Gomar F., Lavie CJ., Perez-Quilis C., Henry BM., Lippi G., “Angiotensin-Converting Enzyme 2 and Antihypertensives (Angiotensin Receptor Blockers and Angiotensin-Converting Enzyme Inhibitors) in Coronavirus Disease 2019,” Mayo Clin Proc., 2020.

[40] Chen Y., Gong X., Wang L., Guo J., “Effects of hypertension, diabetes and coronary heart disease on COVID-19 diseases severity: a systematic review and meta-analysis,” medRxiv, p. doi: https://doi.org/10.1101/2020.03.25.20043133, 2020.

[41] Zuin M., Rigatelli G., Zuliani G., Rigatelli A., Mazza A., Roncon L., “Arterial hypertension and risk of death in patients with COVID-19 infection: Systematic review and meta-analysis,” Journal of Infection, p. doi: https://doi.org/10.1016/j.jinf.2020.03.059, 2020.

[42] Roncon L., Zuin M., Zuliani G., Rigatelli G., “Patients with arterial hypertension and COVID-19 are at higher risk of ICU admission,” British Journal of Anaesthesia, p. doi: https://doi.org/10.1016/j.bja.2020.04.056., 2020.

[43] Phyllis TC., “Hepatitis C Virus-Associated Alterations in Lipid and Lipoprotein Levels: Helpful or Harmful to the Heart,” Clinical Infectious Diseases, vol. 65, pp. 566–567, 2017.

[44] Wei C., Gao Q., Cao Y., Zhong H., et al., “Cholesterol Metabolism Impacts on SARS-CoV-2 Infection Prognosis, Entry, and Antiviral Therapies,” medRxiv, 2020.

[45] Meher G., Bhattacharjy S., Chakraborty H., “Membrane Cholesterol Modulates Oligomeric Status and Peptide-Membrane Interaction of Severe Acute Respiratory Syndrome Coronavirus Fusion Peptide. J Phys Chem B 2019; 123(50): 10654-62.,” The Journal of Physical Chemistry B, vol. 123, no. 50, pp. 10654–10662, 2019.

[46] Ren X., Glende J., Yin J., Schwegmann-Wessels C., Herrler G., “Importance of cholesterol for infection of cells by transmissible gastroenteritis virus,” Virus Research, vol. 137, p. 220–224, 2008.

[47] Nie S., Zhang Z., Zhang Z., et al., “Metabolic disturbances and inflammatory dysfunction predict severity of coronavirus disease 2019 (COVID-19): a retrospective study,” medRxiv, 2020.

[48] Hu X., Chen D., Wu L., He G., Ye W., “Low Serum Cholesterol Level Among Patients with COVID-19 Infection in Wenzhou, China,” SSRN Electron. J., 2020.

[49] Sanjay S., “Statin Drug Therapy May Increase COVID-19 Infection,” Nepalese Medical Journal, vol. 3, no. Epub ahead of print, 2020.

[50] Lin W., Hu L., Zhang Y., Ooi JD., Meng T. bioRxiv, “Single-cell Analysis of ACE2 Expression in Human Kidneys and Bladders Reveals a Potential Route of 2019-nCoV Infection,” bioRxiv, 2020.

[51] Diao B., Feng Z., Wang C., Wang H., Liu L., et al., “Human kidney is a target for novel severe acute respiratory syndrome coronavirus 2 (SARS-CoV-2) infection.,” medRxiv, 2020.

[52] Perico L., Benigni A., Remuzzi G., “Should COVID-19 Concern Nephrologists? Why and to What Extent? The Emerging Impasse of Angiotensin Blockade,” Nephron, 2020.

[53] Alberici F., Delbarba E., Manenti C., Econimo L., Valerio F., “A single center observational study of the clinical characteristics and short-term outcome of 20 kidney transplant patients admitted for SARS-CoV2 pneumonia,” Kidney International, 2020.

[54] Wang J., et al., “COVID-19 in a Kidney Transplant Patient,” Eur Urol, 2020.

[55] Zhou H., Zhang Z., Fan H., Li J., Li M. et al., “Urinalysis, but not blood biochemistry, detects the early renal-impairment in patients with COVID-19,” MedRxiv, 2020.

[56] Cheng Y., Luo R., Wang K., Zhang M., Wang Z., “Kidney disease is associated with in-hospital death of patients with COVID-19,” Kidney International, vol. 97, p. 829–838, 2020.

[57] Avula A., Nalleballe K., Narula N. Sapozhnikov S., Dandu V. et al., “COVID-19 presenting as stroke,” Brain, Behavior, and Immunity, 2020.

[58] Kloka, F.A., Kruipb, M.J.H.A., van der Meerc N.J.M., Arbousd, MS., “Incidence of thrombotic complications in critically ill ICU patients with COVID-19.,” Thrombosis Research, 2020.

[59] Lodigiani C., Iapichino G., Carenzo L., Cecconi M., Ferrazzi P., “Venous and arterial thromboembolic complications in COVID-19 patients admitted to an academic hospital in Milan, Italy,” Thrombosis Research, vol. 191, pp. 9–14, 2020.

[60] Mao L., Jin H., Wang M., Hu Y., Chen S., “Neurologic Manifestations of Hospitalized Patients With Coronavirus Disease 2019 in Wuhan, China,” JAMA Neurology, 2020.

[61] Morelli N., Rota E., Terracciano C., Immovilli P., Spallazzi M. et al., “The Baffling Case of Ischemic Stroke Disappearance from the Casualty Department in the COVID-19 Era,” Eur Neurol, 2020.

[62] Smith CJ., Emsley HC., Gavin CM., Georgiou RF., Vail A., Barberan EM., et al., “Peak plasma interleukin-6 and other peripheral markers of inflammation in the first week of ischaemic stroke correlate with brain infarct volume, stroke severity and long-term outcome.,” BMC Neurol., vol. 4, no. 2, 2004.

[63] Ridker PM., “Anticytokine agents targeting interleukin signaling pathways for the treatment of atherothrombosis.,” Circ Res., vol. 124, no. 3, p. 437–50, 2019.

[64] Oxley TJ., Mocco J., Majidi S., Kellner CP., Shoirah H., “Large-Vessel Stroke as a Presenting Feature of Covid-19 in the Young,” The New England Journal of Medicine, vol. 382, no. 20, 2020.

[65] Hussain A., Bhowmik B., Moreira N., “COVID-19 and diabetes: Knowledge in progress,” Diabetes Research and Clinical Practice, vol. 62, 2020.

[66] Guo W., Li M., Dong Y., et al., “Diabetes is a risk factor for the progression and prognosis of COVID-19,” Diabetes Metab Res Rev., p. e3319, 2020.

[67] Maddaloni E., Buzzetti R., “Covid-19 and diabetes mellitus: unveiling the interaction of two pandemics,” Diabetes Metabolism research review, 2020.

[68] Moujaess E., Kourie HR., Ghosn M., “Cancer patients and research during COVID-19 pandemic: A systematic review of current evidence,” Critical Reviews in Oncology / Hematology, vol. 150, 2020.

[69] Xia Y., Jin R., Zhao J., Li W., Shen H., “Risk of COVID-19 for patients with cancer,” Lancet Oncology, vol. 21, 2020.

